# Spatiotemporal Evolution of SARS-CoV-2 Alpha and Delta Variants during a Large Nationwide Outbreak in Vietnam, 2021

**DOI:** 10.1101/2023.01.03.22283384

**Authors:** Nguyen Thi Tam, Nguyen To Anh, Trinh Son Tung, Pham Ngoc Thach, Nguyen Thanh Dung, Van Dinh Trang, Le Manh Hung, Trinh Cong Dien, Nghiem My Ngoc, Le Van Duyet, Phan Manh Cuong, Hoang Vu Mai Phuong, Pham Quang Thai, Nguyen Le Nhu Tung, Dinh Nguyen Huy Man, Nguyen Thanh Phong, Vo Minh Quang, Pham Thi Ngoc Thoa, Nguyen Thanh Truong, Tran Nguyen Phuong Thao, Dao Phuong Linh, Ngo Tan Tai, Ho The Bao, Vo Trong Vuong, Huynh Thi Kim Nhung, Phan Nu Dieu Hong, Le Thi Phuoc Hanh, Le Thanh Chung, Nguyen Thi Thanh Nhan, Ton That Thanh, Do Thai Hung, Huynh Kim Mai, Trinh Hoang Long, Nguyen Thu Trang, Nguyen Thi Hong Thuong, Nguyen Thi Thu Hong, Le Nguyen Truc Nhu, Nguyen Thi Han Ny, Cao Thu Thuy, Le Kim Thanh, Lam Anh Nguyet, Le Thi Quynh Mai, Tang Chi Thuong, Le Hong Nga, Tran Tan Thanh, Guy Thwaites, H. Rogier van Doorn, Nguyen Van Vinh Chau, Thomas Kesteman, Le Van Tan, OUCRU COVID-19 research group

## Abstract

In 2021, Vietnam experienced a large nationwide outbreak of COVID-19, with over 1.7 million infections and 32,000 deaths. We generated 1,303 SARS-CoV-2 whole-genome sequences, and mapped out the public health measures alongside the evolutionary trajectory of the pathogen. The Alpha variant caused sporadic outbreaks in early 2021 prior to the upsurge in cases associated with the Delta variant from June onward. The Delta variant outbreak was almost entirely confined to the AY.57 lineage, accounting for 99.2% of 1,212 Delta variant sequences, and resulting from a single introduction. Viral dispersal from the north, where it was first introduced into Vietnam, to the south, marked the start of the nationwide outbreak, with the south subsequently seeding the virus back to the other regions. Distinct AY.57 phylogenetic clusters in different regions of Vietnam were documented, pointing to the impact of in-country lockdown. Genomic surveillance is critical to inform pandemic response.

## INTRODUCTION

SARS-CoV-2 Alpha and Delta variants of concern (VoC) were first detected in November 2020 in England and India, respectively. Both variants carry key mutations (including N501Y, P681H and H69/V70 deletion for the Alpha variant, and L452R, P681R and T478K for the Delta variant) in the spike protein that make the viruses bind more efficiently to the angiotensin-converting–enzyme 2 receptor of human cells and/or escape antibody neutralisation induced by infection or vaccination with ancestral strain [1-3]. Because of these features, the Alpha variant is estimated to be 50% more transmissible than the ancestral strain, and the Delta variant is approximately 40-60% more transmissible than the Alpha variant [4]. As a consequence, both variants rapidly spread around the world and became the dominant viruses responsible for two consecutive COVID-19 waves in 2021.

Having reported only 1,465 PCR-confirmed cases of SARS-CoV-2 infection and 35 deaths in 2020 [5], Vietnam was one of the few countries to successfully control community transmission of SARS-CoV-2 during the first year of the pandemic. The cornerstone of this success was the early introduction of border and travel restrictions, meticulous contact tracing coupled with mass testing and strict quarantine, and intensive mass communication to the public [5, 6]. However, the importations of the highly transmissible Alpha variant followed by the Delta variant in early 2021 resulted in a large nationwide outbreak, with over 1.7 million infections and 32,000 deaths reported by the end of 2021 [7].

Genomic surveillance has been one of the top priorities of the WHO. In addition to enabling early detection of new SARS-CoV-2 variants, it has provided significant insights into the evolutionary trajectories of SARS-CoV-2 in time and space [8], critical to inform pandemic response. Yet, most of the studies published about genetic evolution of SARS-CoV-2 are based on datasets from high-income countries with relatively open borders [9-11], and little is known about the transmission dynamics of SARS-CoV-2 in settings with extremely limited number of viral importations such as Vietnam. Here, we focused our analysis on spatiotemporal evolution of SARS-CoV-2 Alpha and Delta variants in Vietnam in 2021. Our aim was also to map out the patterns of viral evolution and diffusion against the public health measures implemented in Vietnam as the pandemic progressed during the study period.

## MATERIALS AND METHODS

### Vietnam COVID-19 containment approach in 2021

After successfully controlling community transmission during the first eight months in 2020, Vietnam cautiously reopened its borders, allowing some repatriation flights, while steadily lifting in-country travel restrictions [5]. However, following the report of new SARS-CoV-2 variants in the UK and India in late 2020, now known as Alpha and Delta variants, respectively, Vietnam suspended all inbound flights from countries reporting the detection of community transmission associated with these two variants. Additionally, all travelers entering Vietnam were subjected to 14-day isolation, and testing on the day of arrival and on day 14 of quarantine. Yet, after 28 days of no community transmission, on 18^th^ January 2021, two clusters of SARS-CoV-2 infection with epidemiological links were detected in two neighboring provinces in the north of Vietnam, Hai Duong and Quang Ninh [12]. This was followed by the detection of several other community transmission clusters of unclear origin in several provinces across the country in subsequent months before the start of the 2021 major outbreak (Figure 1). At the national level, the responses were initially targeted e.g., lockdown at community/province/city level where a community transmission cluster was detected. However, in response to the escalation of the outbreak starting in July 2021 [7], Vietnam suspended all arriving international flights in May 2021, and re-applied in-country travel restriction (Figure 1) until 10^th^ October 2021 when nearly 70% of the eligible population had received at least one dose of vaccine [7]. In parallel, under the zero COVID-19 policy, Vietnam implemented the meticulous contact tracing and mass testing of the cases and their contacts conducted by provincial Centers for Diseases Control (CDC). This had enabled accurate identification of cases of community transmission origin alongside the demographic data for analysis [13].

**Figure 1:**
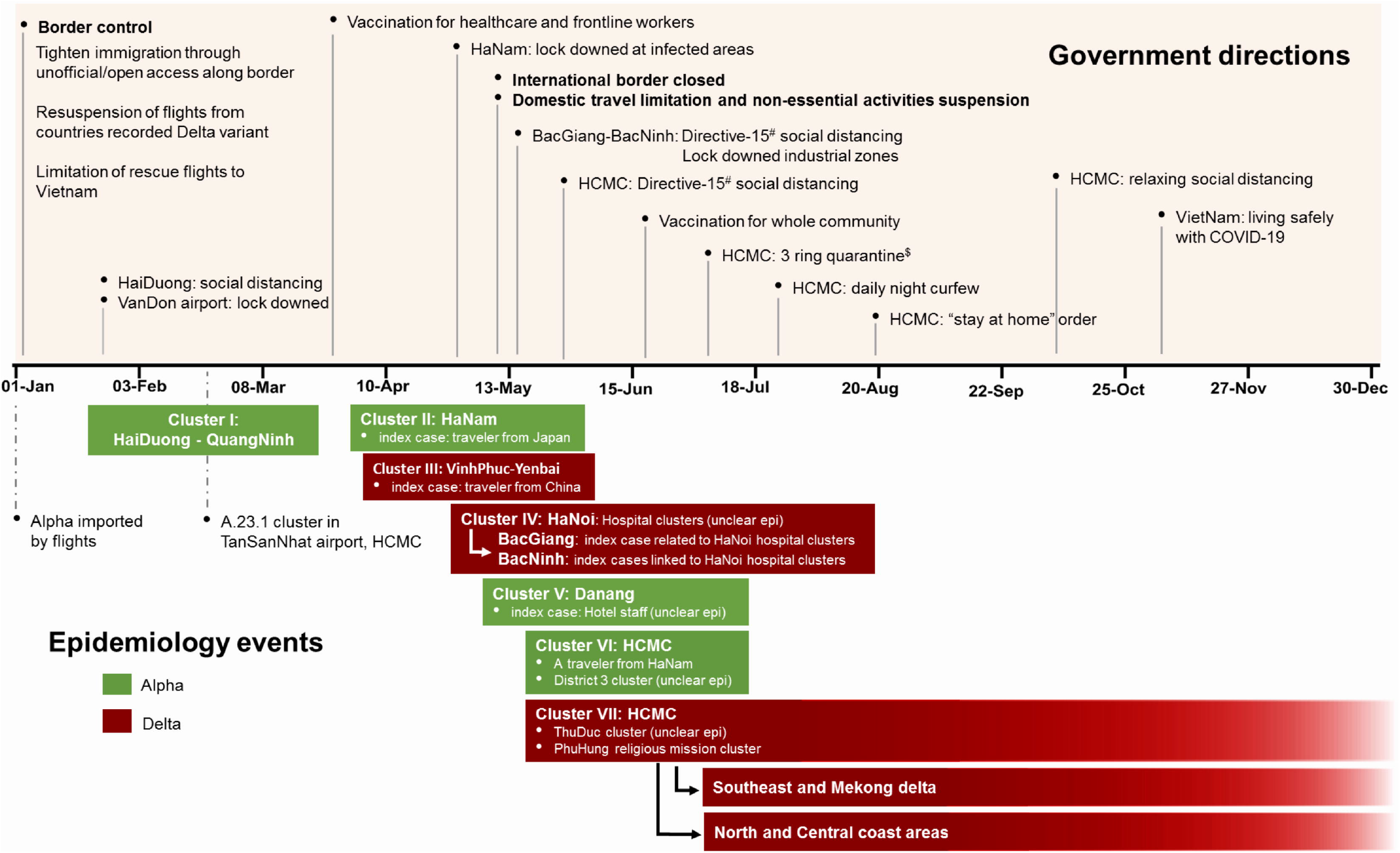
Government directions and COVID-19 epidemiology events in Vietnam in 2021. **Note to Figure 1**: ^#^Directive-15: Suspension of non-essential services/businesses and mass gatherings, applying physical distance of 2 meters when contact with others, banning the gatherings of 20 people or more in one place and 10 people or more outside workplaces and limitation of movements. ^$^3 ring quarantine: apply Directive-16 whole city (bans gatherings of two more people in public and asks people to only leave homes for emergencies, food, medicine, work in factories, and businesses that involve essential goods and services), lockdown at residential areas with covid-19 case report, home quarantine for F1. HCMC: Ho Chi Minh city

### Settings and sampling approach

The present study was conducted at the National Hospital for Tropical Diseases (NHTD) in Hanoi, and the Hospital for Tropical Diseases (HTD) in Ho Chi Minh City, Vietnam. Both NHTD and HTD are tertiary referral hospitals for COVID-19 patients in the north and south of Vietnam, respectively. The laboratories of these hospitals were responsible for SARS-CoV-2 diagnosis and sequencing in Vietnam. Therefore, nasopharyngeal swab (NPS) samples submitted to NHTD and HTD laboratories for testing and sequencing were either from provincial CDC or from inpatients being treated at these two hospitals.

To increase the chance of successfully obtaining the virus genome, we first applied a preselection criterion based on the cycle threshold (Ct) value of the tested samples generated by the Lightmix Modular SARS-CoV-2 RdRp/E gene assay (Tib Molbiol, Berlin, Germany) [14]. Accordingly, at NHTD, only NPS samples with a cycle threshold (Ct) value of ≤30 for the RdRp gene were eligible, while at HTD, a sample with Ct value of ≤25 for the E gene was included. Additionally, because of the availability of the resources, between January and June 2021 when community transmission remained limited, our approach focused on new community clusters detected through contact tracing under zero-COVID strategy. Between July and December 2021 during which community transmission was escalating, the selection of samples for sequencing was carried out using WHO recommendations [15]. Epidemiological data, and data on infections and deaths were retrieved from e-hospital record or provided by the National Institute of Hygiene and Epidemiology and the Vietnamese Ministry of Health. Here, we focused our analysis on sequences obtained cases of locally acquired infection, and generated by NHTD or HTD laboratories, of which we had accurate sampling date and demographic data.

### RNA extraction, whole genome sequencing and sequence assembly

Total RNA was extracted from NPS using QIAamp viral RNA mini kit (Qiagen, Hilden, Germany) following the manufacturer’s instructions, and finally eluted in 50μl of elution buffer (provided with the extraction kit). Whole genome amplification was performed on the extracted RNA using either the long pooled amplicons protocol, developed by the University of Sydney [16, 17], or the ARTIC V3 protocol [18] on an Illumina MiSeq platform as previously described. Library preparation was carried out using the Nextera XT Library preparation kit (Illumina, USA), followed by library quantification using KAPA Library Quant Kit (Kapa Biosystems, Wilmington, MA, USA), according to the manufacturer’s instructions. The prepared library was sequenced using MiSeq reagent kit V2 (300 cycles) on a Miseq platform (Illumina). For each run, tested samples were multiplexed and differentiated by double indexes using IDT-ILMN Nextera DNA UD indexes (IDT).

Sequence assembly of the obtained sequencing data was carried out using a reference-based mapping approach available in CLC genomics workbench (v.21.0.4) and Geneious 8.1.5 (Biomatters, San Francisco, CA, USA). This method involved mapping of sequencing output of individual samples to a reference genome (WuHan-Hu-1: NC_045512, Alpha: EPI_ISl_905782, Delta: EPI_ISL_1942165), followed by manual editing of the obtained consensus, as described previously [16]. The consensus sequences generated in this study were submitted to the National Center for Biotechnology Information under the assigned accession numbers ON458864-ON459533, ON459545-ON459608, ON755375-ON755859 and OP647358-OP647411.

### Lineage classification, sequences alignment and recombination detection

SARS-CoV-2 lineage classification of the obtained consensuses was determined using PANGO lineage [19] with pangolin v4.1.2 and pangolin-data v1.13 [20]. The analysis was carried out using the Ultrafast Sample Placement on Existing Trees option to assign the lineage based on the nearest lineage on existing global tree. Sequence alignment was carried out using tool available on Nextclade [21] and Minimap2 [22]. Recombination detection was inferred using a combination of Freyja v 1.3.10 [23] and sc2rf [24].

### Phylogenetic analysis

To explore the phylogenetic relationship of Vietnamese Alpha and Delta variant sequences, we focused our analysis on complete coding region (29,408 bp) of the obtained sequences, and applied maximum likelihood (ML) method using TIM2 nucleotide substitution model with invariant for Alpha variant, and UNREST+F0+R4 for Delta variant as suggested by IQ tree [25]. Support for individual nodes was assessed using a bootstrap procedure with 1000 replicates. To assess the placement of the Vietnamese variants in the context of global sequences, we employed the phylogenetic framework incorporated in NextClade [21], taking into account representatives of global sequences submitted to GISAID.

### Phylogeography and phylodynamics of Delta variant AY.57 lineage

To study the phylogeographics and phylodynamics of the pathogen, we applied Bayesian phylogenetic interference in BEAST v1.10.4 [26]. We first excluded identical sequences and sequences of low quality (e.g., internal gaps). We then used TempEst 1.5 to assess the temporal signal of the dataset [27]. Subsequently, we excluded the sequences not conforming to a linear evolutionary pattern as suggested by TempEst. For phylogeographic analysis, we divided Vietnam into eight major geographic regions (Figure 5). We employed a Bayesian Markov chain Monte Carlo framework (available in BEAST) with 1 billion steps and sampling every 100,000 steps using the general time reversible (GTR) nucleotide substitution model with invariant (as suggested by IQ-TREE to be the best-fit model) under an uncorrelated relaxed clock model [28], and a Bayesian skygride coalescent tree prior (10 groups) [29]. We assessed convergence using Tracer version 1.5 [30]. We selected a burn-in threshold of 10% and accepted effective sample size values above 200. Maximum-clade credibility (MCC) tree was then summarized with TreeAnnotator (available in the BEAST package) and visualized in Figtree version 1.4.2 [31].

To assess the effective population size trajectory, we applied Bayesian Skyride model using the above frame work. Finally, we used codon-based method (HyPhy) available in MEGA5 to measure the selection pressure on coding sequences of the pathogen genome, by estimating the ratio of nonsynonymous to synonymous substitution (dN/dS) [32].

### Ethics

The study received approvals from the Institutional Review Board of the Hospital for Tropical Diseases in Ho Chi Minh City, Vietnam and the Oxford Tropical Research Ethics Committee. Our study formed part of the national COVID-19 response, and used deidentified throat swabs from COVID-19 patients. Thus, the need for individual informed consent was waived.

## RESULTS

### COVID-19 situation in Vietnam in 2021

The COVID-19 situation in Vietnam in 2021 was characterized by the two distinct phases. The first one was between January and April, during which a total of 1,632 infections and no deaths were recorded. This was followed by a second phase occurring between May and December, with a large upsurge in the number of infections (n=1,727,398) and deaths (n=32,359) (Figure 2A).

**Figure 2:**
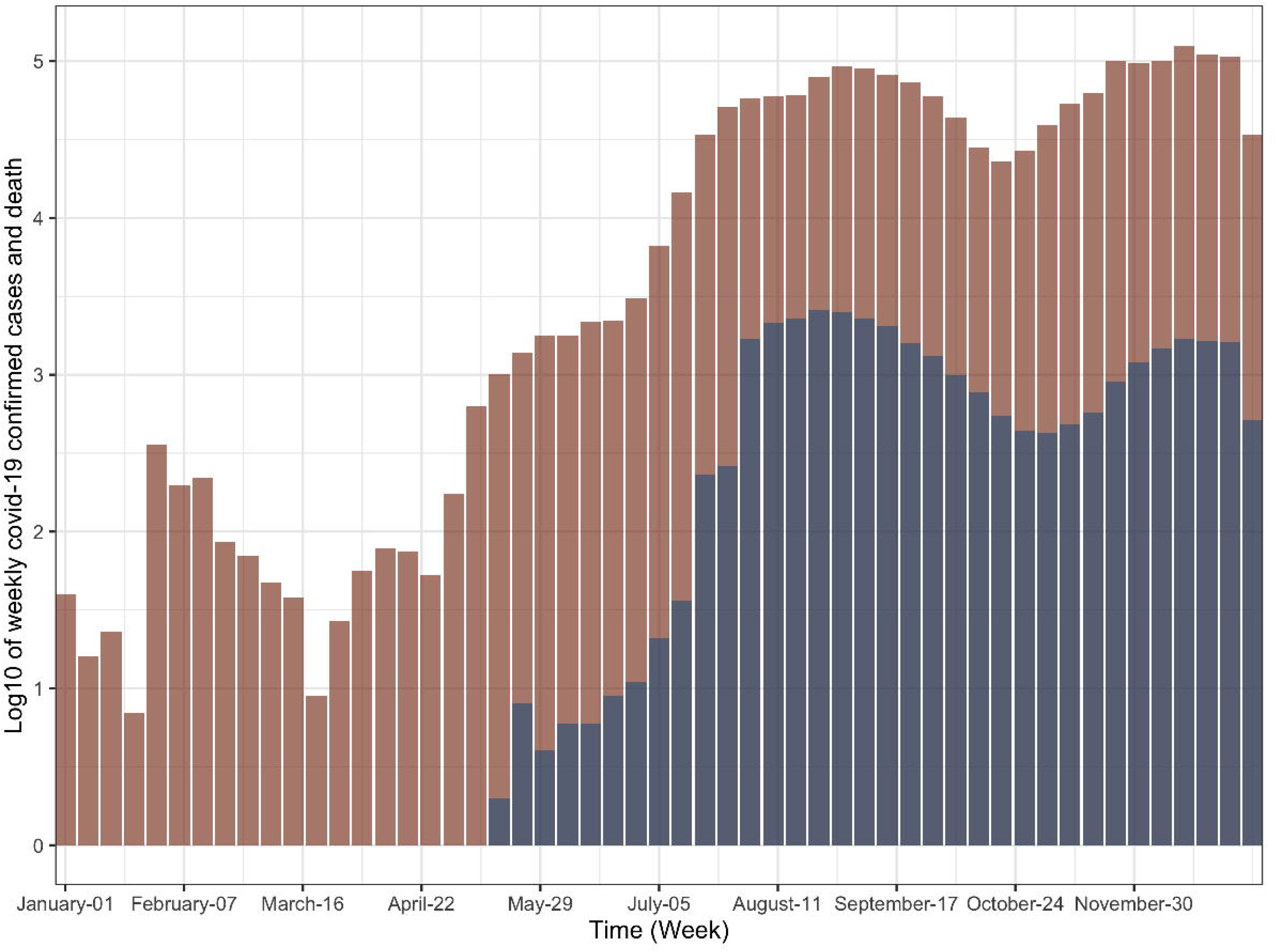
A) Bar chart showing the number infections (brown bars) and deaths (grey bars) during the 2021 outbreak in Vietnam, B) monthly number of SARS-CoV-2 variants detected among cases of community transmission between January and December 2021. Delta variant non-AY.57 lineage includes B.1.617.2 (n=1), AY.23 (n=3), AY.79 (n=3), AY.85 (n=1), AY.6 (n=1), and AY.38 (n=1). Others include lineages B.1.637 (n=2) and A.23.1 (n=5).

#### Results of whole genome sequencing and the study subjects

In total, 1,365 NPS were selected for WGS. Eventually, 1,303 genome sequences were successfully obtained (Figure 3). No recombinants were detected. The majority belonged to Delta variant (n=1,222, 93.8%), followed by Alpha (n= 74, 5.7%), A.23.1 (n=5, 0.4%), and B.1.637 (n=2, 0.2%) variants. Of Delta sequences, 1,212 (99.2%) were assigned to AY.57 lineage, and the remaining was assigned to AY.23 and AY.79 (n=3 each), AY.6, AY.38, AY.85, and B.1.617.2 (n=1 each).

**Figure 3:**
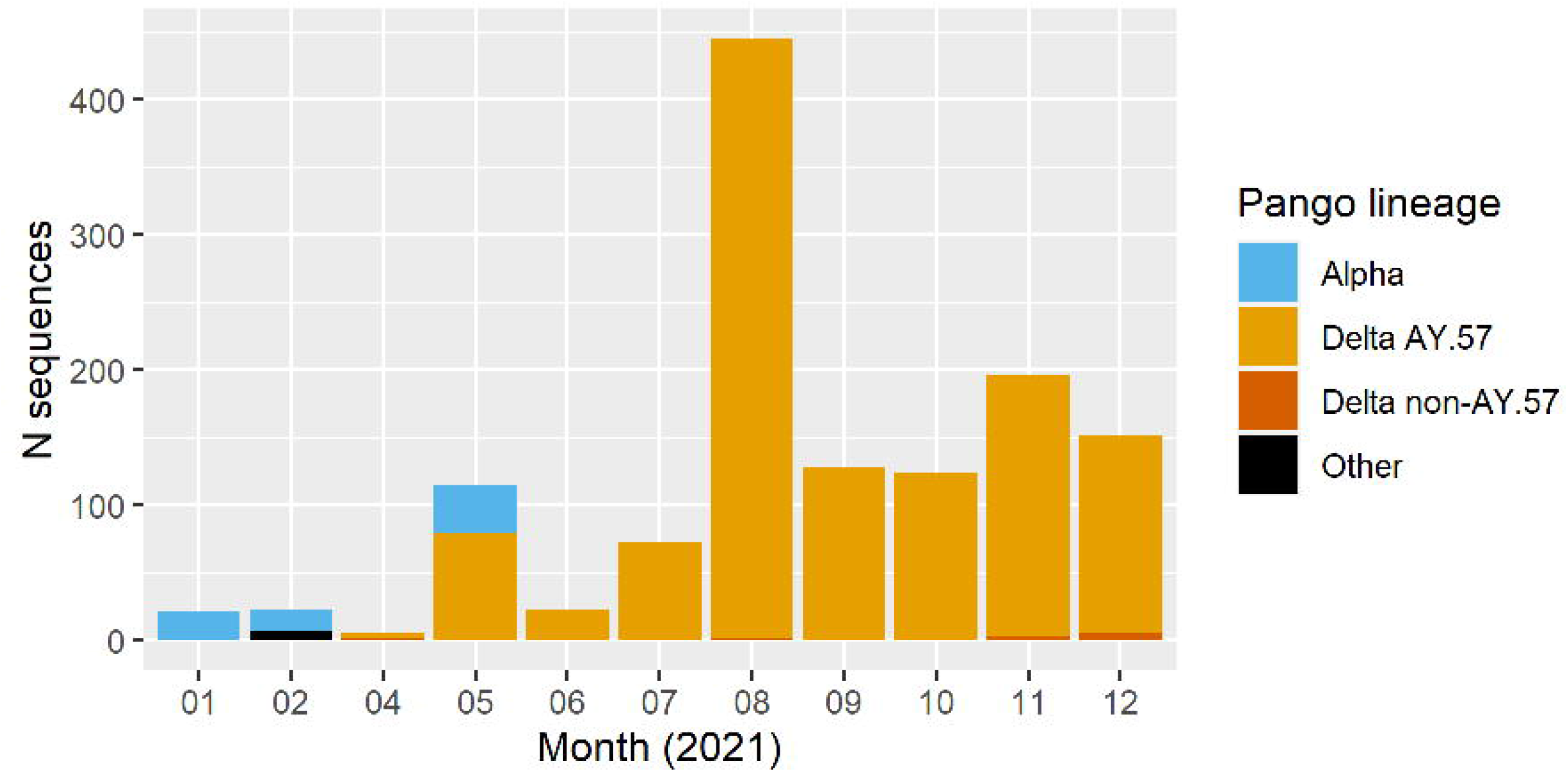
Workflow of the study

Temporally, the obtained sequences distributed throughout the year, with sequences of the Alpha and A.23.1 documented between January and May 2021. The first three Delta variant sequences, including two AY.57 and one B.1.617.2, were detected late April 2021 (Figure 2B). From June onward, Delta variant was the only variant detected, coinciding with the upsurge in the number of infections and deaths of the 2021 outbreak (Figure 2A).

The demographic features of the study subjects are presented in Table 1. The characteristics of the A.23.1 variant infected cases were previously reported [12].

**Table 1.** Demographics of the study participants

|  | Number of sequences<br>(N=1303) | Alpha<br>(N = 74) | AY.57<br>(N = 1212) |
| --- | --- | --- | --- |
| **Age (Years)** |  |  |  |
| Median (Q1-Q3) | 43 (29 - 61) | 35 (30-55) | 44 (29 - 61) |
| Missing | 38 (2.9%) | 31 (41.9%) | 2 (0.2%) |
| **Gender** |  |  |  |
| Female | 689 (52.9%) | 22 (29.7%) | 661 (54.5%) |
| Male | 578 (44.4%) | 21 (28.4%) | 551 (45.5%) |
| Missing | 36 (2.8%) | 31 (41.9%) |  |
| **Region** |  |  |  |
| Centre | 120 (9.2%) | 24 (32.4%) | 94 (7.8%) |
| North | 504 (38.7%) | 44 (59.5%) | 450 (37.1%) |
| South | 679 (52.1%) | 6 (8.1%) | 668 (55.1%) |

#### Phylogenetic relatedness between sequences of Alpha variant

Phylogenetic analysis of Alpha variant sequences showed that they clustered into four major groups (Figure 4 and Supplementary Figure 1), corresponding to the sporadic community transmission clusters detected in the north, central, and south of Vietnam in early 2021 (Figure 1). This suggested that multiple introductions of Alpha variant into Vietnam occurred between January and May 2021. However, community wide transmission was not sustained, likely attributable to the rapid and rigorous domestic responses (Figure 1 and Figure 2A).

**Figure 4:**
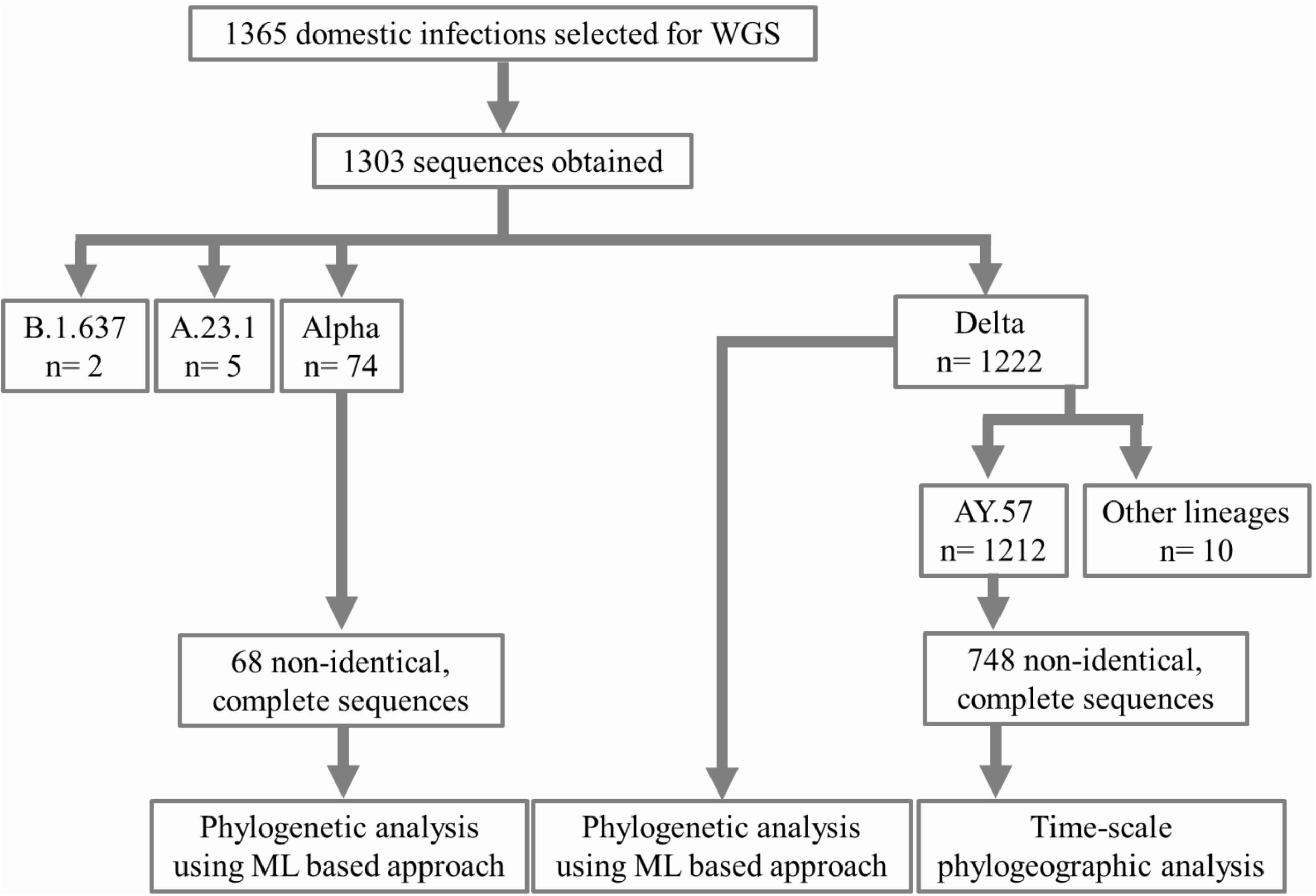
Maximum likelihood tree of complete coding sequences of Alpha variant circulating in Vietnam in 2021. Phylogenetic clusters were named accordingly to community clusters recorded during the study period shown in Figure 1.

#### Phylogenetic relatedness between sequences of Delta variant

The reconstructed phylogenetic trees derived from 1,222 Delta variant sequences confirmed the dominance of AY.57 lineage (99.2%) detected in Vietnam throughout 2021, with a small number of the sequences grouped into non-AY.57 groups (Supplementary Figure 2). This was consistent with the results of pangolin lineage classification.

#### Spatiotemporal evolution of Delta variant AY.57 lineage in Vietnam

Because Delta variant AY.57 lineage was the dominant virus detected during the study period, we focused our phylogeographic analysis on 1,212 sequences of this lineage detected among cases of community transmission. After removing identical sequences, sequences of low quality, and outliers as suggested by TempEst, a total of 748 non-identical sequences were available for analysis. TempEst analysis indicated a positive correlation between genetic divergence and sampling time; the retrieved R^2^ value of 0.4538 (F = 438.4117, p value < 0.001), suggesting a moderate temporal signal in the included sequences (Supplementary Figure 3). Results of phylogeographic analysis confirmed that AY.57 lineage responsible for the first nationwide wave of COVID-19 in Vietnam in 2021 was first introduced into the Northeast region in early 2021 (Figure 5) and the estimated time to the most recent common ancestor (tMRCA) was March 14, 2021 (95%CI: February 22, 2021 to April 8, 2021). This coincided with the detection of a community transmission cluster in Vinh Phuc and Yen Bai provinces linked with travelers in the north in April 2021 (Figure 1). In the following months, the northeast and Red river delta regions then acted as a source seeding the virus to other neighboring regions, and especially to provinces in the southeastern region. However, viral dispersal between provinces/cities was not frequently observed (Figure 5).

**Figure 5:**
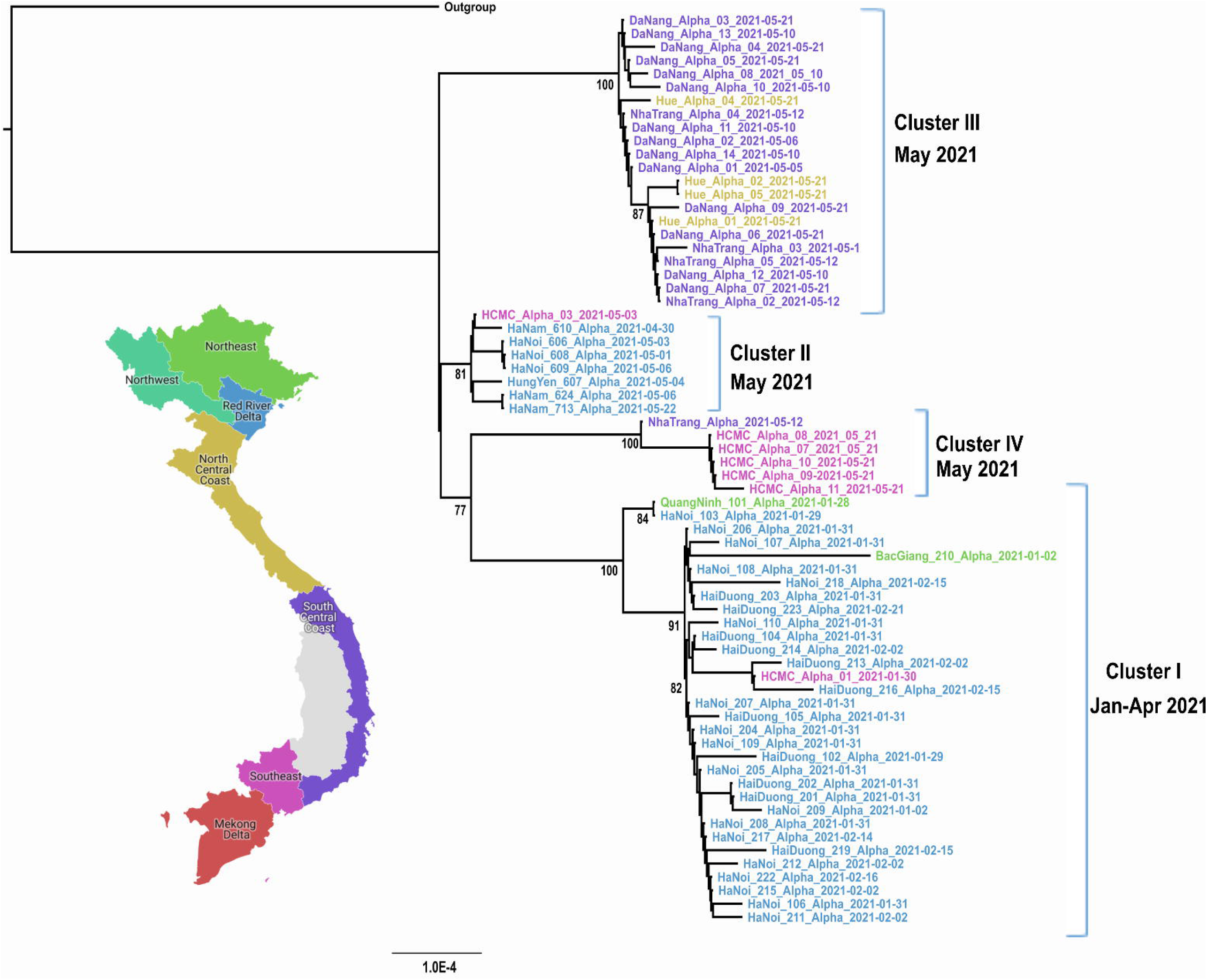
Maximum clade credibility trees illustrating the phylogeography of AY.57 lineage in Vietnam in 2021. Branches are color-coded according to locations of sampling. Posterior probabilities >0.8% and state probabilities >75% (black circles) are indicated at all nodes

Our analysis also revealed that between July and December 2021, the southeastern region was the main source seeding the virus back to the north and the rest of Vietnam. Additionally, we observed the establishment of multiple localized clusters of AY.57 lineage elsewhere in the south central coastal region and the Red river delta (Figure 5).

#### Phylodynamics of Delta variant lineage AY.57

Bayesian Skyride plot reflecting the dynamics of effective population size of AY.57 lineage circulating within Vietnam is shown in Figure 6. Although the results should be interpreted with caution, we observed a sharp increase in genetic diversity of the pathogen between April and August 2021 (Figure 6A), reflecting the expansion of the AY.57 lineage across the country, coinciding with the start of the large nationwide outbreak from May onward (Figure 1 and Figure 2A). In the following months, the viral population size remained relative stable, despite some remarkable fluctuations in the number of viral sequences obtained (Figure 6B). This was followed by a slight decrease in the genetic diversity of the pathogen in the end of November.

**Figure 6:**
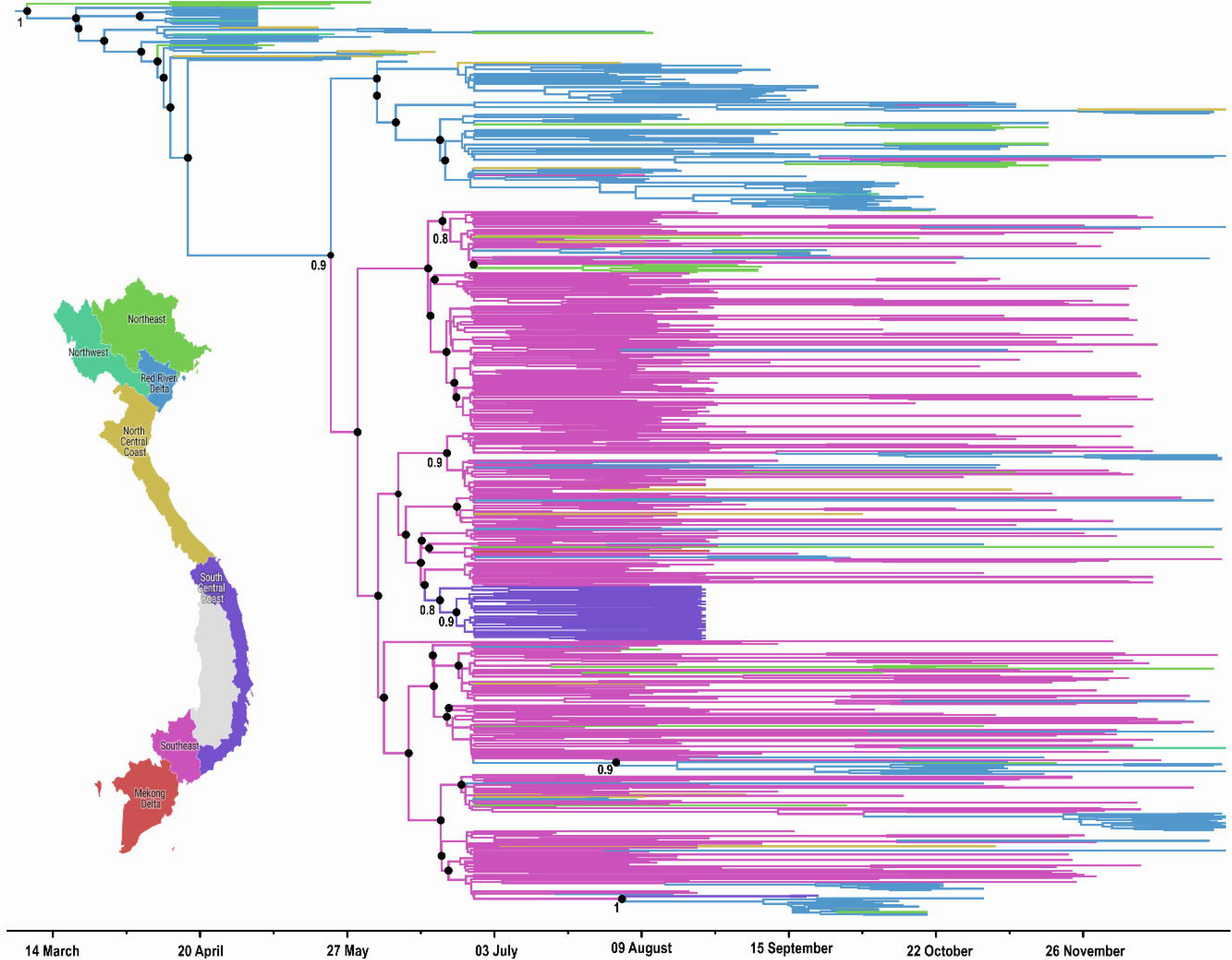
**A)** Bayesian skyride plot illustrating the relative genetic diversity of AY.57 lineage in Vietnam during the year 2021, and B) the Number of AY.57 sequences used for analysis. Blue shading (Panel A) indicates 95% highest posterior density interval.

#### Natural selection

The estimated mean of dN/dS value of polyprotein coding sequences was 0.86. The dN/dS values for each individual protein coding gene were as follows: Open Reading Frame (ORF) 1a: 0.69, ORF 1b: 1.45, spike protein coding sequence: 0.59, ORF 3a: 1.44, envelope protein coding sequence: 1.14, Matrix protein coding sequence: 0.27, ORF: 0.86, ORF 7a: 6; 1.3, ORF 7b: 0.82, ORF 8: 0.37, and N protein coding sequence: 0.53. Finally, our estimated evolutionary rate of AY.57 coding sequences from Vietnamese community cases sampled in 2021 was estimated to be 5.29×10^−4^ (95%CI: 4.966×10^−4^-5.639×10^−4^) substitutions per site per year.

## DISCUSSION

We report the evolutionary history of SARS-CoV-2 variants and the potential impact of public health measures on the spread of the virus during an explosive nationwide outbreak in Vietnam in 2021. The Alpha variant was first introduced into Vietnam in early 2021, shortly after it was first discovered in the UK in November 2020, via different importation events as suggested by the results of phylogenetic analysis. However, we only documented sporadic clusters of community transmission associated with the Alpha variant until May before the Delta variant took over from June onward. This was likely attributable to the rigorous containment approach applied in Vietnam at the time, also known as zero COVID-19 policy, and the transmissibility of the Alpha variant itself. Notably, during the same period the Alpha variant was responsible for a new wave of COVID-19 elsewhere, including in the UK and in Europe [33, 34].

Unlike the Alpha variant, the Delta variant was not detected in Vietnam until April 2021. Despite rigorous public health measures applied during the next two months, the virus rapidly spread to other provinces in the north, where it was first detected, and in the south before it caused an explosive nationwide outbreak from May onward. Our genomic surveillance data revealed that AY.57 lineage of the Delta variant was the responsible virus with a tMRCA dating back in March 2021.

Viruses of the Delta variant have been divided into over 240 sublineages [35]. Of these, AY.57 lineage is characterized by the presence of signature mutations in ORF1a (P380S, P1640L, A3209V, V3718A, and T3750I), and in spike protein (A222V). The origin of AY.57 lineage causing the outbreak in Vietnam remains to be determined. However, the result of tMRCA analysis suggested that this lineage had emerged shortly after the discovery of the Delta variant in India in November 2020. We may however underestimated the tMRCA of the AY.57 lineage since we only sequenced 1212 AY.57 sequences, while over 1.7 million cases were reported during the study period. Collectively, the data emphasize the importance of active genomic surveillance to inform pandemic responses.

In spite of the rapid expansion of the AY.57 lineage and the escalation of the outbreak from July onward, our phylogeographic analysis suggested that viral spread between localities in Vietnam only sporadically occurred. Of note, between July and September, Vietnam implemented strict national lockdown with very limited in-country travel, and further tightened border controls. This may have influenced the dispersal of the virus and explained the establishment of distinct phylogenetic clusters of AY.57 lineage in different geographic locations across Vietnam. Notably, in other countries, such as the UK and the US, where travel restriction and border control were less stringent as compared to those applied in Vietnam, viral dispersal across the localities was more apparent [36, 37]. Additionally, during the study period, 198 and 195 sublineages of the Delta variant were documented in the UK and the US, respectively [35]. In contrast, the 2021 outbreak in Vietnam was almost entirely confined to the AY.57 lineage, likely via a single introduction as suggested by the tight cluster of the Vietnamese AY.57 lineage on the NextClade-based global phylogenetic tree.

We were able to reveal important insights in the evolutionary dynamics of the AY.57 lineage. Accordingly, we showed that there was a sharp increase in relative genetic diversity of the pathogen between April and July 2021, just around the start of the nationwide outbreak. This suggested that the transmission into the community, followed by uncontrolled spread for the first time in Vietnam, was likely attributable to the high transmissibility of the Delta variant. This had resulted in the natural expansion in genetic diversity of the pathogen as it spread among a susceptible population. Notably, between April and July 2021, only a very small proportion of Vietnamese people were vaccinated against COVID-19 (51.000 (0.0005%) and 6.2 million (0.06%) by 1 April and 31 July of a population of 97 million, respectively) [38]. Therefore, at the time Vietnamese people remained largely naïve to SARS-CoV-2 infection [7]. Collectively, our data therefore reflect the natural evolutionary dynamics of AY.57 lineage.

Our estimated dN/dS ratio for entire coding region suggested that the evolution of AY.57 lineage was driven by a weak purifying selection, although this varied across the virus genome, with ORF1b, ORF3a, E gene and ORF7a exhibiting a positive selection. Of note, a recent report showed that the vaccines had played a role in selective adaptation of SARS-CoV-2 Delta variant [39]. As a consequence, different mutation profiles were recorded between vaccine breakthrough infection and non-breakthrough infection groups [39]. The role of vaccination in shaping the evolution of SARS-CoV-2 merits further research. Finally, our estimated evolutionary rate of AY.57 lineage was within the range of previously estimated values for the Delta variant [36, 40].

Our study has some limitations. We acknowledged a potential bias towards our referral-hospital based sampling approach, especially from July 2021 onwards, when moderate and asymptomatic cases started to be managed at home. As a consequence, we cannot exclude the possibility of the co-circulation of other SARS-CoV-2 variants in Vietnam, which was not captured by our sampling approach. However, this was very unlikely given the tight control border implemented in Vietnam during the study period, and our results are consistent with sequences uploaded on GISAID [35].

In summary, we report how the Alpha and Delta variants were introduced, spread, and evolved during the large nationwide outbreak of COVID-19 in Vietnam in 2021. Rigorous public health measures successfully contained the Alpha variant during the first wave. The introduction of the highly transmissible Delta variant AY.57 in an immunologically naïve population however resulted in an explosive nationwide outbreak, coinciding with a rapid increase in genetic diversity of the pathogen. Distinct clusters of AY.57 lineage were established in different geographic locations, pointing to the impact of public health containment on the evolution and the diffusion of SARS-CoV-2. Our study thus emphasizes how genomic surveillance is critical to inform pandemic response.

## Data Availability

All data produced in the present study are available upon reasonable request to the authors

## ACKNOWLEDGEMENTS

We would like to thank to Dr.Le Nguyen Minh Hoa and technicians at Department of Microbiology and Molecular, National Hospital for Tropical Diseases for collecting swab samples and initial testing of SARS-CoV-2 diagnostic. We are grateful to OUCRU team for their supporting for whole genome sequencing and data entry. We thank the submitters from many laboratories for the sequences on the GISAID database.

The genomic surveillance work was funded by Wellcome (222574/Z/21/Z). LVT and GT are supported by the Wellcome Trust of Great Britain (204904/Z/16/Z and 106680/B/14/Z, respectively).

OUCRU COVID-19 research group: Chambers Mary, Choisy Marc, Dong Huu Khanh Trinh, Dong Thi Hoai Tam, Du Hong Duc, Dung Vu Tien Viet, Fisher Jaom, Flower Barney, Geskus Ronald, Hang Vu Thi Kim, Ho Quang Chanh, Ho Thi Bich Hai, Ho Van Hien, Hung Vu Bao, Huong Dang Thao, Huynh le Anh Huy, Huynh Ngan Ha, Huynh Trung Trieu, Huynh Xuan Yen, Kestelyn Evelyne, Kesteman Thomas, Lam Anh Nguyet, Lawson Katrina, Leigh Jones, Le Kim Thanh, Le Dinh Van Khoa, Le Thanh Hoang Nhat, Le Van Tan, Lewycka Sonia Odette, Lam Minh Yen, Le Nguyen Truc Nhu, Le Thi Hoang Lan, Nam Vinh Nguyen, Ngo Thi Hoa, Nguyen Bao Tran, Nguyen Duc Manh, Nguyen Hoang Yen, Nguyen Le Thao My, Nguyen Minh Nguyet, Nguyen To Anh, Nguyen Thanh Ha, Nguyen Than Ha Quyen, Nguyen Thanh Ngoc, Nguyen Thanh Thuy Nhien, Nguyen Thi Han Ny, Nguyen Thi Hong Thuong, Nguyen Thi Hong Yen, Nguyen Thi Huyen Trang, Nguyen Thi Kim Ngoc, Nguyen Thi Kim Tuyen, Nguyen Thi Ngoc Diep, Nguyen Thi Phuong Dung, Nguyen Thi Tam, Nguyen Thi Thu Hong, Nguyen Thu Trang, Nguyen Thuy Thuong Thuong, Nguyen Xuan Truong, Nhung Doan Phuong, Ninh Thi Thanh Van, Ong Phuc Thinh, Pham Ngoc Thanh, Phan Nguyen Quoc Khanh, Phung Ho Thi Kim, Phung Khanh Lam, Phung Le Kim Yen, Phung Tran Huy Nhat, Rahman Motiur, Thuong Nguyen Thi Huyen, Thwaites Guy, Thwaites Louise, Tran Bang Huyen, Tran Dong Thai Han, Tran Kim Van Anh, Tran Minh Hien, Tran Phuong Thao, Tran Tan Thanh, Tran Thi Bich Ngoc, Tran Thi Hang, Tran Tinh Hien, Trinh Son Tung, van Doorn H. Rogier, Van Nuil Jennifer, Vidaillac Celine Pascale, Vu Thi Ngoc Bich, Vu Thi Ty Hang, Yacoub Sophie. HTD COVID-19 research group: Nguyen Van Vinh Chau, Nguyen Thanh Dung, Le Manh Hung, Huynh Thi Loan, Nguyen Thanh Truong, Nguyen Thanh Phong, Dinh Nguyen Huy Man, Nguyen Van Hao, Duong Bich Thuy, Nghiem My Ngoc, Nguyen Phu Huong Lan, Pham Thi Ngoc Thoa, Tran Nguyen Phuong Thao, Tran Thi Lan Phuong, Le Thi Tam Uyen, Tran Thi Thanh Tam, Bui Thi Ton That, Huynh Kim Nhung, Ngo Tan Tai, Tran Nguyen Hoang Tu, Vo Trong Vuong, Dinh Thi Bich Ty, Le Thi Dung, Thai Lam Uyen, Nguyen Thi My Tien, Ho Thi Thu Thao, Nguyen Ngoc Thao, Huynh Ngoc Thien Vuong, Huynh Trung Trieu, Pham Ngoc Phuong Thao, Phan Minh Phuong. EOCRU COVID-19 research group: Bachtiar, Andy, Baird, Kevin J., Dewi, Fitri, Dien, Ragil, Djaafara, Bimandra A., Elyazar, Iqbal E., Hamers, Raph L., Handayani, Winahyu, Kurniawan, Livia Nathania, Limato, Ralalicia, Natasha, Cindy, Nuraeni, Nunung, Puspatriani, Khairunisa, Rahadjani, Mutia, Rimainar, Atika, Saraswati, Shankar, Anuraj H., Suhendra, Henry, Sutrisni, Ida Ayu, Suwarti, Tarino, Nicolas, Timoria, Diana, Wulandari, Fitri. OUCRU-NP COVID-19 research group: Basnyat Buddha, Duwal Manish, Gautum Amit, Karkey Abhilasha, Kharel Niharika, Pandey Aakriti, Rijal Samita, Shrestha Suchita, Thapa Pratibha, Udas Summita.

## ABOUT THE AUTHOR

Dr. Nguyen Thi Tam is a Postdoc Scientist at Oxford University Clinical Research Unit, Hanoi, Vietnam. She is a key person of genomic surveillance of SARS-CoV-2 variants emerging in Vietnam since 2020. She also interested in situation and molecular mechanisms of AMR of bacterial pathogens in Vietnam.

---

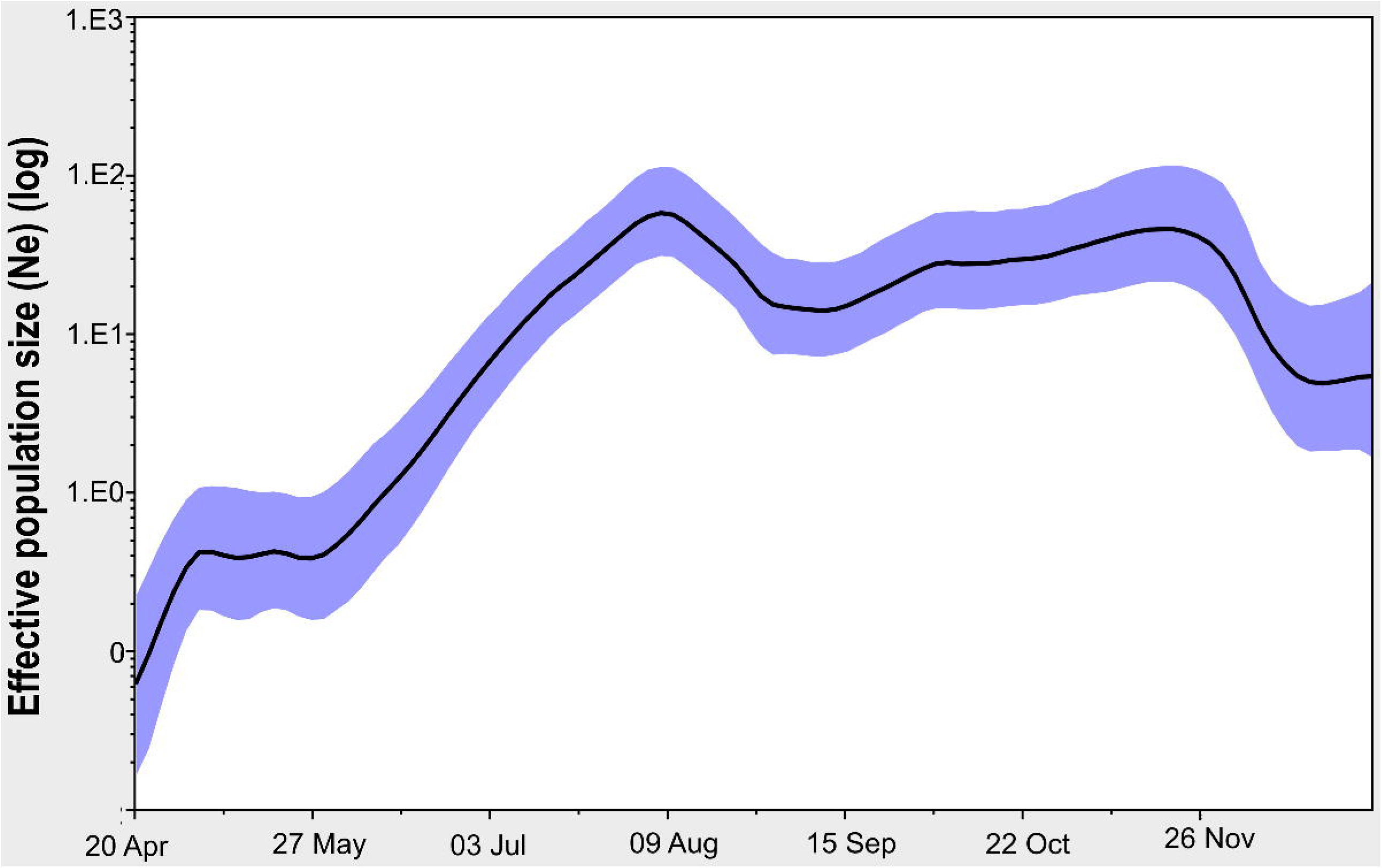

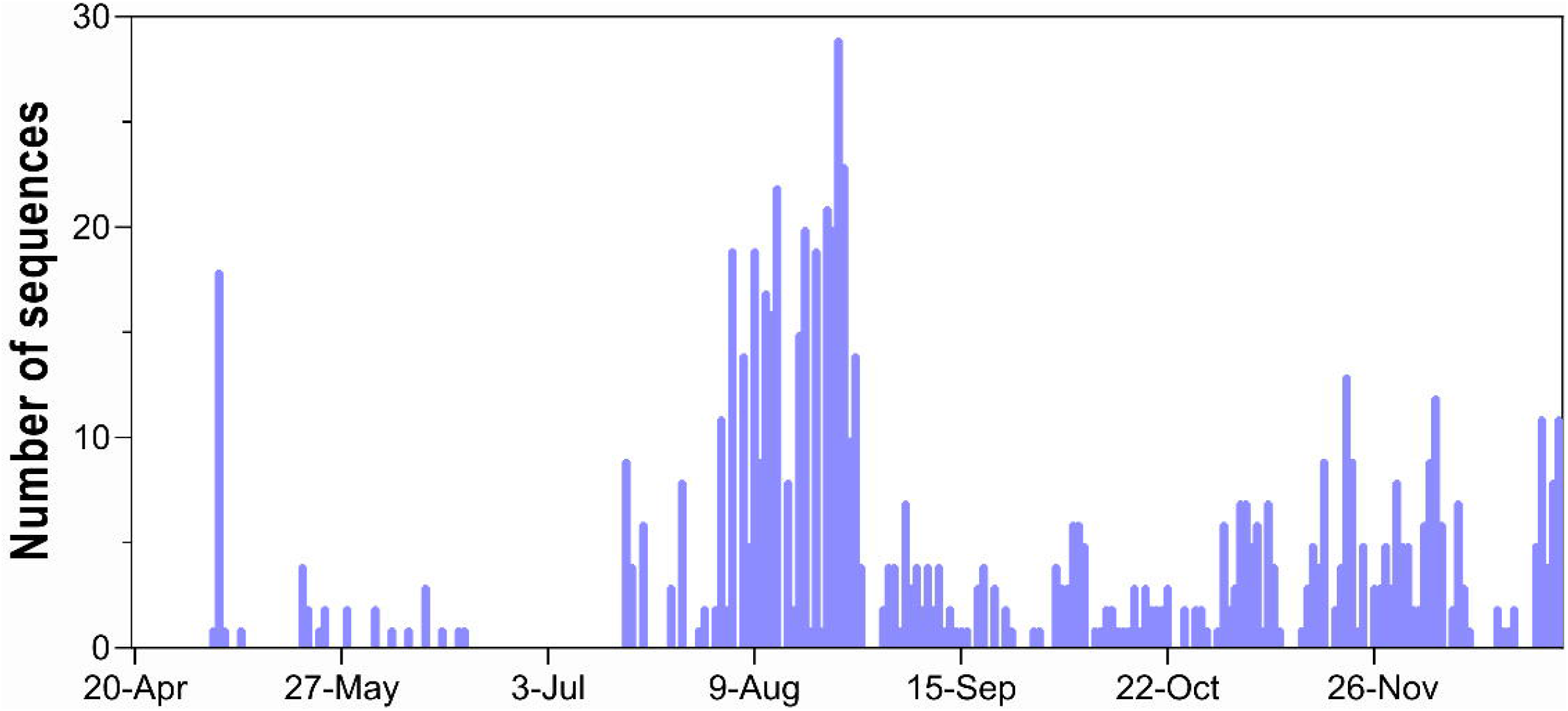

