## Supplementary material for "Spatiotemporal Evolution of SARS-CoV-2 Alpha and Delta Variants during a Large Nationwide Outbreak in Vietnam, 2021"

### SUPPLEMENTARY MATERIALS

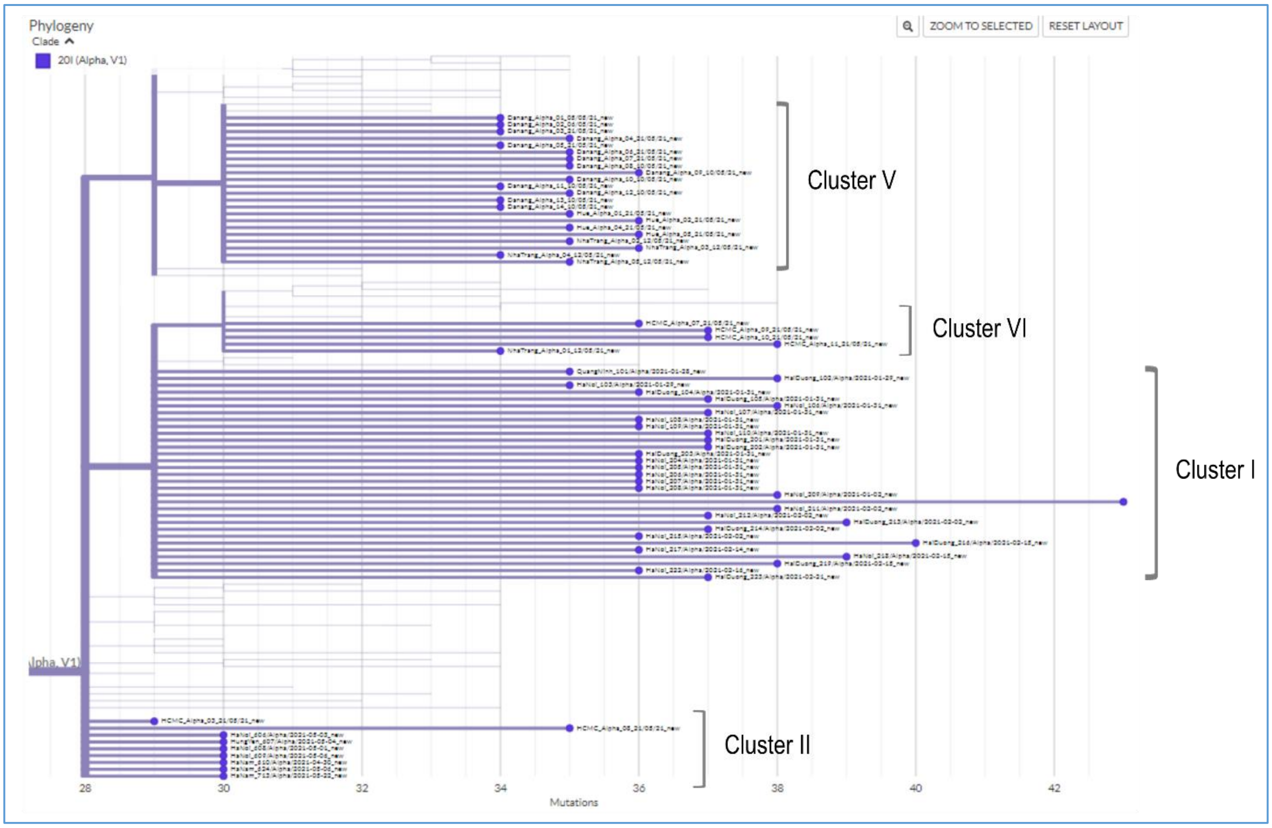


**Supplementary Figure 1:** NextClade generated phylogenetic tree demonstrating the relatedness between the Vietnamese Alpha variant sequences and global sequences submitted to GISIAD. Phylogenetic clusters were named accordingly to community clusters recorded during the study period shown in Figure 1.

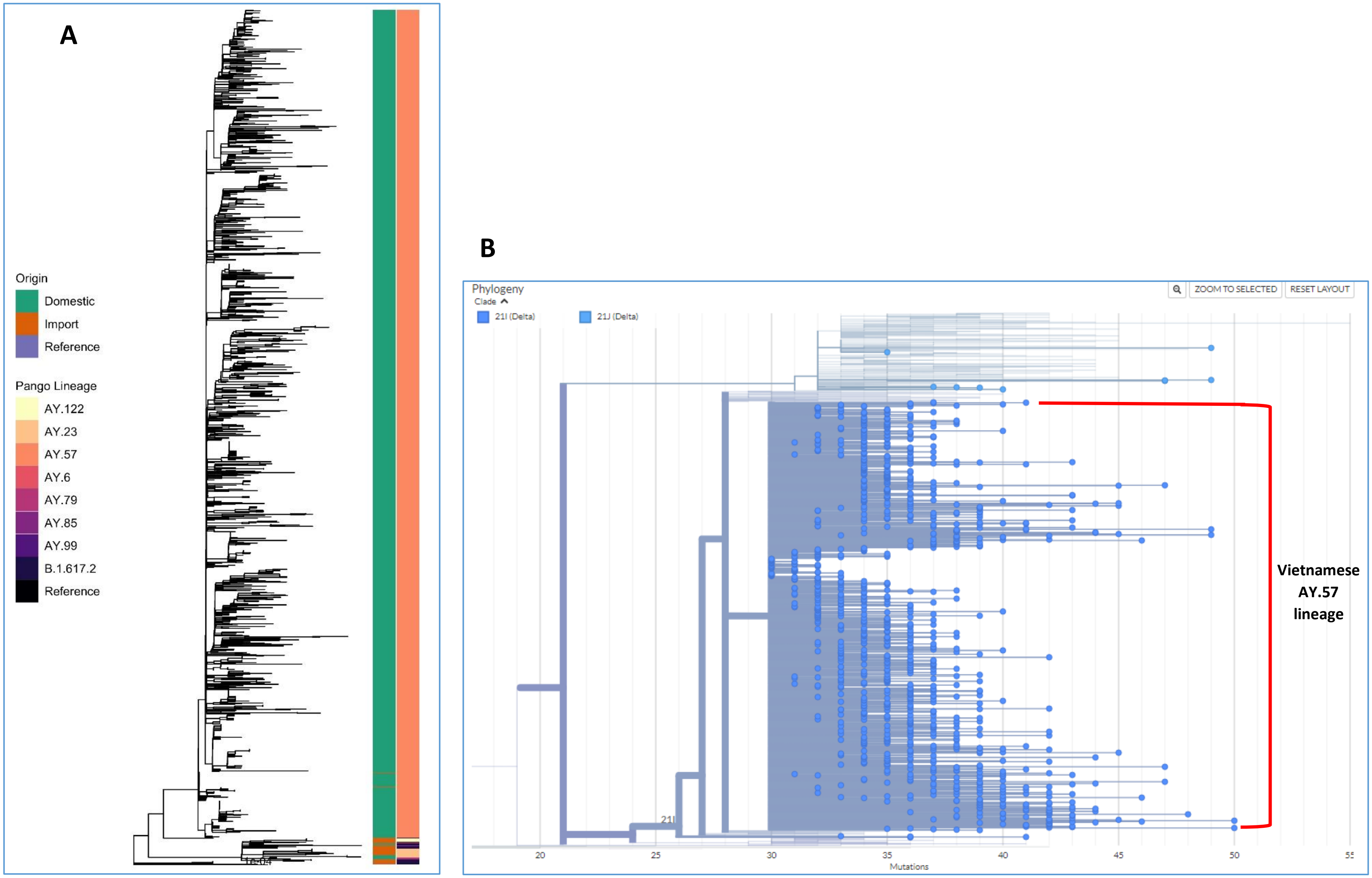


**Supplementary Figure 2: A)** Reconstructed ML tree depicting the relationship between Delta variants detected in Vietnam, and **B)** NexClade Based phylogenetic analysis illustrating the placement of the Vietnamese sequences among global sequences submitted GISAID

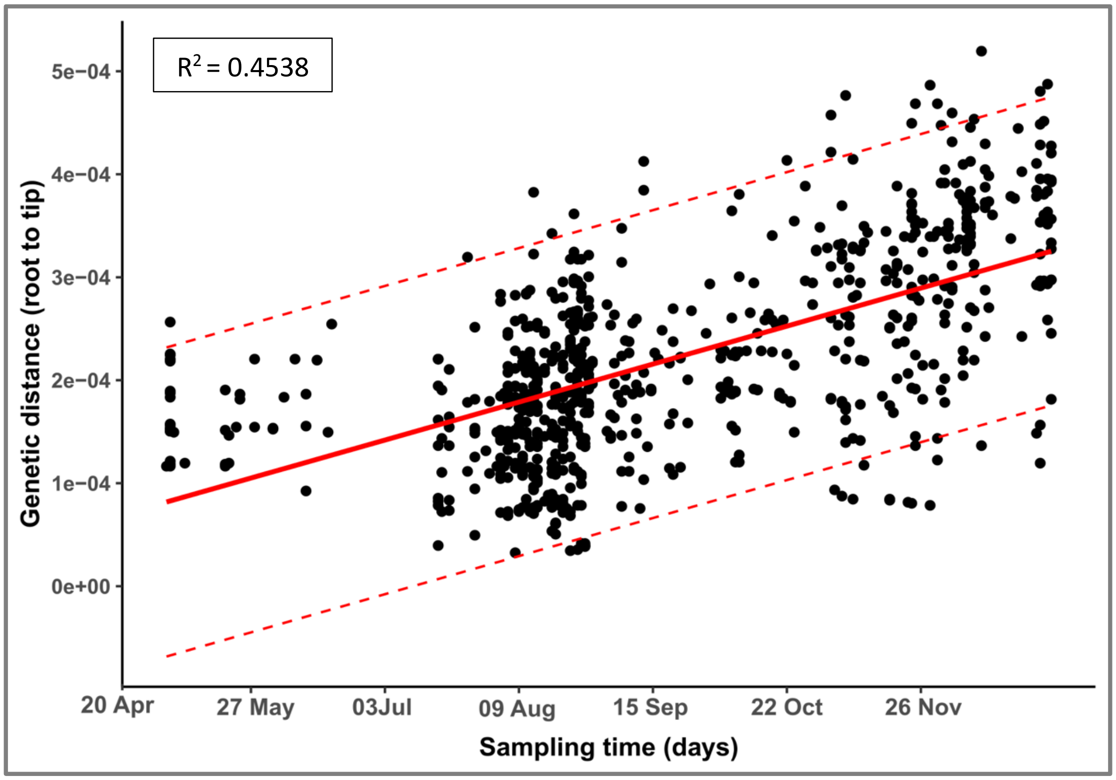


**Supplementary Figure 3:** Root to tip regression of AY.57 coding sequences utilizing for the evolution analysis. The solid line indicates the regression line; the dotted lines represent upper and lower limits of 95% confidence interval. Outliers were excluded from subsequence spatiotemporal evolutionary analysis.
